# Non-specific Primers Reveal False-negative Risk in Detection of COVID-19

**DOI:** 10.1101/2020.04.07.20056804

**Authors:** Wei Liu

## Abstract

**Background:** A novel coronavirus disease 2019 (COVID-19) broke out in Wuhan of Hubei province and had spread throughout the world since December 2019. Because the clinically diagnosed cases in Hubei province were reported for the first time on February 13, 2020, a very high peak of new cases in China was observed. The reason why so many clinically diagnosed cases appeared was not clear.

**Methods:** All data of new cases in China were acquired from WHO situation reports. Linear fitting was used to infer the ability to detect COVID-19 infections. Primer-BLAST and nucleotide blast were applied to check the specificity of primers. Expression data of human mRNA in different tissues was obtained from Human Protein Atlas.

**Findings:** Based on the data and analysis of changes of new laboratory-confirmed cases and new clinically diagnosed cases, it was inferred that there were many false-negative results in all clinically diagnosed cases in Hubei province. There were eight non-specific primers in dozens of primers used in clinical or research detection of COVID-19. Among them, a pair of primer for the ORF1ab regions of SARS-CoV-2 genome well matched some human mRNAs such as Cathepsin C transcripts. Compared to other transcripts, Cathepsin C mRNA had a high abundance in tonsil, lung and small intestine.

**Interpretation:** Some non-specific RT-PCR primers could cause the serious interference during RT-PCR amplification so as to increase the risk of false-negative diagnoses for COVID-19 infections.

**Funding:** Key Research Project of the Higher Education of Henan Province

**Research in context:** *Evidence before this study:* The author searched PubMed on April 15, 2020, for papers that describe false-negative RT-PCR detection of COVID-19 by using the search terms “COVID-19”, “false-negative” and “RT-PCR”, with no language or time restrictions. Eleven investigations only presented the rate of false-negative detection or the importance of positive chest CT finding. There were no reports referring the primer problems of false-negative detection in COVID-19 infections.

*Added value of this study:* The author had found that some primers could amplify the human mRNA in specimens, which mixed SARS-CoV-2 viral particles and other tissue cells. A pair of primer provided by China CDC could vastly match the sequences of human *CTSC* transcripts with high abundance. That could lead to false-negative results in detection of COVID-19 infections.

*Implications of all the available evidence:* Although there were so many false-negative results in detection of COVID-19 infections in China, the exact reason was not clear. Problems in sampling and test condition were discussed thoroughly, but conclusions were usually contradictory. Therefore, the work could promote the verification of the false-negative detection of COVID-19 infections in China.

## Introduction

The World Health Organization (WHO) had declared coronavirus disease 2019 (COVID-19) a pandemic on March 11, 2020, in which SARS-CoV-2 virus had spread around the world.^1^ SARS-CoV-2 virus was first isolated from patients in Wuhan and identified as a novel coronavirus in late December, 2019.^2^ It caused an outbreak of COVID-19 in China in late January, 2020. Although so far WHO had reported more than 82,000 confirmed coronavirus infections in China, there was few new native cases in late March, 2020. However, there were more than 2 million confirmed coronavirus infections around world.^3^ As an easy and rapid method to diagnose coronavirus virus, including SARS-CoV-2, SARS and so on, reverse transcription-polymerase chain reaction (RT-PCR) was commonly used to confirm COVID-19 infections worldwide. Some institutes or universities provided several RT-PCR assays to diagnose 2019-nCoV (previous name of COVID-19) and the protocols.^4^

Because of occurrence of abnormal results, several articles from Chinese research groups had referred to the RT-PCR diagnoses for COVID-19 infections according to China CDC guideline.^5^ False-negative RT-PCR test results had been reported by a previous study.^6^ Five in 167 patients had presented negative results in first RT-PCR detection for 2019 novel coronavirus with positive chest CT finding. All of them were finally confirmed with COVID-19 infection by second, third or multiple repeated RT-PCR detections. A correspondence on viral load in upper respiratory specimens showed that some patients received negative or positive results alternately and repeatedly by using the primers targeted the ORF1ab region of novel coronavirus genome,^7^ and similar situation was also described in another correspondence.^8^ A recent letter discussed positive RT-PCR test results after two consecutively negative results in four patients recovered from COVID-19, and could not rule out false-negative results.^9^

The investigation had paid attention to the above phenomenon and tried to understand the extent of false-negative results from changes of new cases of the COVID-19 infections in China. Based on the primers provided by research institutes from different countries, especially primers from China, detailed analysis of non-specificity of primer sequences had been conducted, and interference of human mRNA targeted by the primer was discussed deeply for RT-PCR detection of COVID-19 infections.

## Methods

### Data source

Numbers of new cases of COVID-19 infections were released by WHO.^3^ Data spanned from January 22, 2020 to March 6, 2020. Seventeen specific primer pairs for detection of Covid-19, as shown in Table S1, were provided by several institutes or universities from different countries.^4^ Among them, two sets of primers, including forward primer (F), reverse primer (R) and fluorescence probe, were designed by National Institute for Viral Disease Control and Prevention (IVDC), China CDC.^5^ Since the onset of the COVID-19 outbreak, two sets of primers had recommended to guide disease prevention and control in China.^10,11^ All mRNA expressions in different human organs were obtained from Human Protein Atlas (HPA, https://www.proteinatlas.org/).^12^

### Analysis of primer specificity

Primer-BLAST (https://www.ncbi.nlm.nih.gov/tools/primer-blast/index.cgi) was applied to determine whether specific primers for SARS-CoV-2 virus show significant match with human RNA or not.^13^ Two “Exclusion” options should be selected to avoid PREDICTED results, and other options were set as default. Nucleotide blast (https://blast.ncbi.nlm.nih.gov/Blast.cgi) was also used to check the specificity of primers, in which options Refseq RNA was chosen as database, word size was set to 7, and expect threshold was set to 1000.^14^

## Results

Figure 1 showed the new cases of the COVID-19 infections in China from January 22 to March 6, 2020. Blue, green and red lines represented laboratory-confirmed cases, clinically diagnosed cases and total new cases, respectively. Numbers of new laboratory-confirmed cases and new clinically diagnosed was not reported from February 17 to February 19, 2020 and clinically diagnosed cases in Hubei province was removed by WHO from February 20, 2020. Except for positive results in RT-PCR detection, clinically diagnosed patients met all clinical diagnostic conditions including chest CT images.^15, 16^ Since China reported 12,289 clinically diagnosed cases in Hubei province for the first time, number of total new cases grew explosively on February 13, 2020, whereas number of clinically diagnosed cases decreased dramatically to 888 from February 14 to February 16, 2020. Meanwhile number of new laboratory-confirmed cases decreased gradually from less than 2,000 to about 1,000, which were obviously less than 3,893 new laboratory-confirmed cases on February 5, 2020. The inset of Figure 1 showed the linear fitting of the number of laboratory-confirmed cases to the number of days from January 22 to February 5, 2020. The predicted number of detection could reach about 5512 according to linear fitting formula (the predicted number of detection = 260.32 × days - 475.44) on February 13, 2020, while there were 1820 laboratory-confirmed cases.

**Figure 1:**
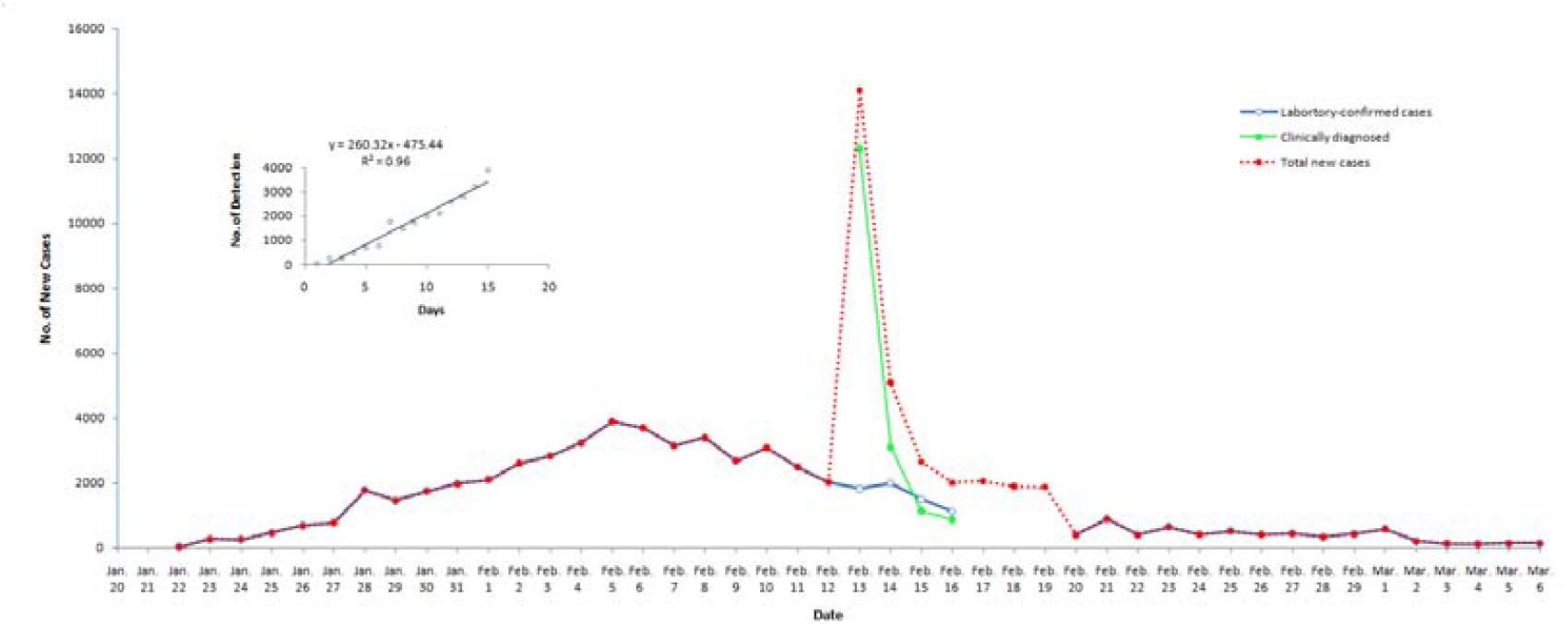
New cases of the Covid-19 infections in China from January 22 to March 6, 2020. Inset showed the number of detection vs. days from January 22 to February 5, 2020. Linear fitting formula was as follows: *y* = 260.32 *x* - 475.44.

Figure 2 showed the position of all primer pairs on the genome of SARS-CoV-2 Wuhan-Hu-1 strain. There were seventeen primer pairs, thirteen for universal RT-PCT and four for two sets of nested RT-PCR. The lengths of all primers were between 17 and 26. The numbers of primer pairs targeted ORF1ab, spike protein S, envelop protein E and nucleoprotein N nucleotide sequences were 7, 2, 1 and 7, respectively. Three primer pairs overlap to some extent in *RdRp* gene in ORF1ab region, while two and three primer pairs overlap to some degree in nucleoprotein N gene, respectively. Except for nested RT-PCR with amplicon size of 346 to 547 nt, the lengths of all RT-PCR amplification products were between 57 and 158 nt.

**Figure 2:**
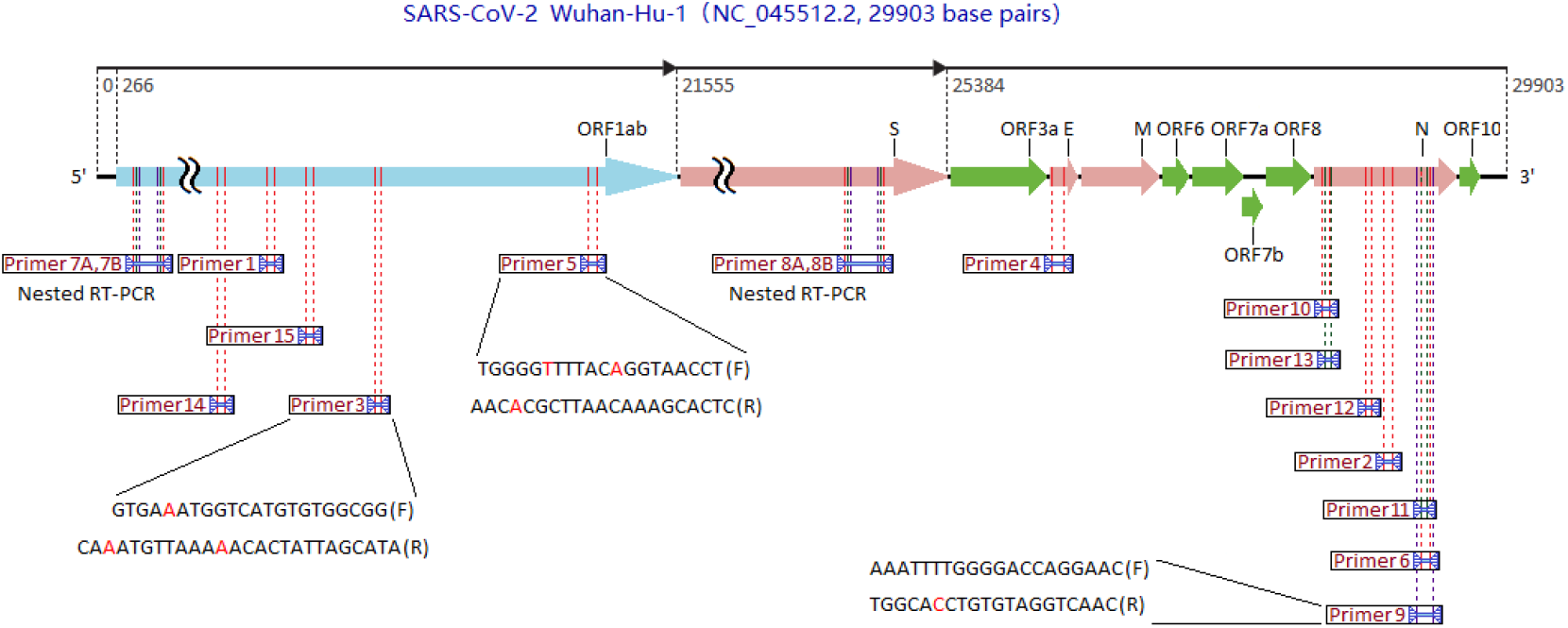
Schematic diagram of prime positions located on the SARS-CoV-2 genome. Bases in red color were determined according to SARS-CoV-2 Wuhan-Hu-1 sequence. A portion of the respective length of ORF1ab and S was omitted.

The specificity of all primers was checked with Primer-BLAST and nucleotide blast to ascertain whether it were a primer for one or more human RNA transcripts or not. It is important to note that more meaningful matches could be obtained using Primer-BLAST than nucleotide Blast, and choosing Refseq mRNA database could acquire a little more matches than choosing Refseq RNA database with Primer-BLAST.

Table 1 listed the non-specific primers originally designed for SARS-CoV-2 and their unintended human mRNA targets found by using Primer-BLAST. Besides those non-specific primers, all other primers in Figure 2 were also checked with Primer-BLAST. Three new combinations of the forward and reverse sequences of primer 1 (for ORF1ab regions) matched many human mRNA transcripts to a higher degree, whereas there were no any primer pair matching human mRNAs for primer 2 (for N gene). Cross combinations of the forward and reverse sequences of primer 1 and 2 did not match more human mRNAs than the above combinations. Several consecutive nucleotides at 3’ ends of four new primer pairs composed of the reverse sequence of primer 1 perfectly matched those of *CTSC, ZNF7, FYCO1* and *RRAGB* transcripts, and its probe (5’-FAM-CCGTCTGCGGTATGTGGAAAGGTTATGG-BHQ1-3’) could not match *CTSC, ZNF7, FYCO1* and *RRAGB* transcripts more than nine nucleotides.

**Table 1:**
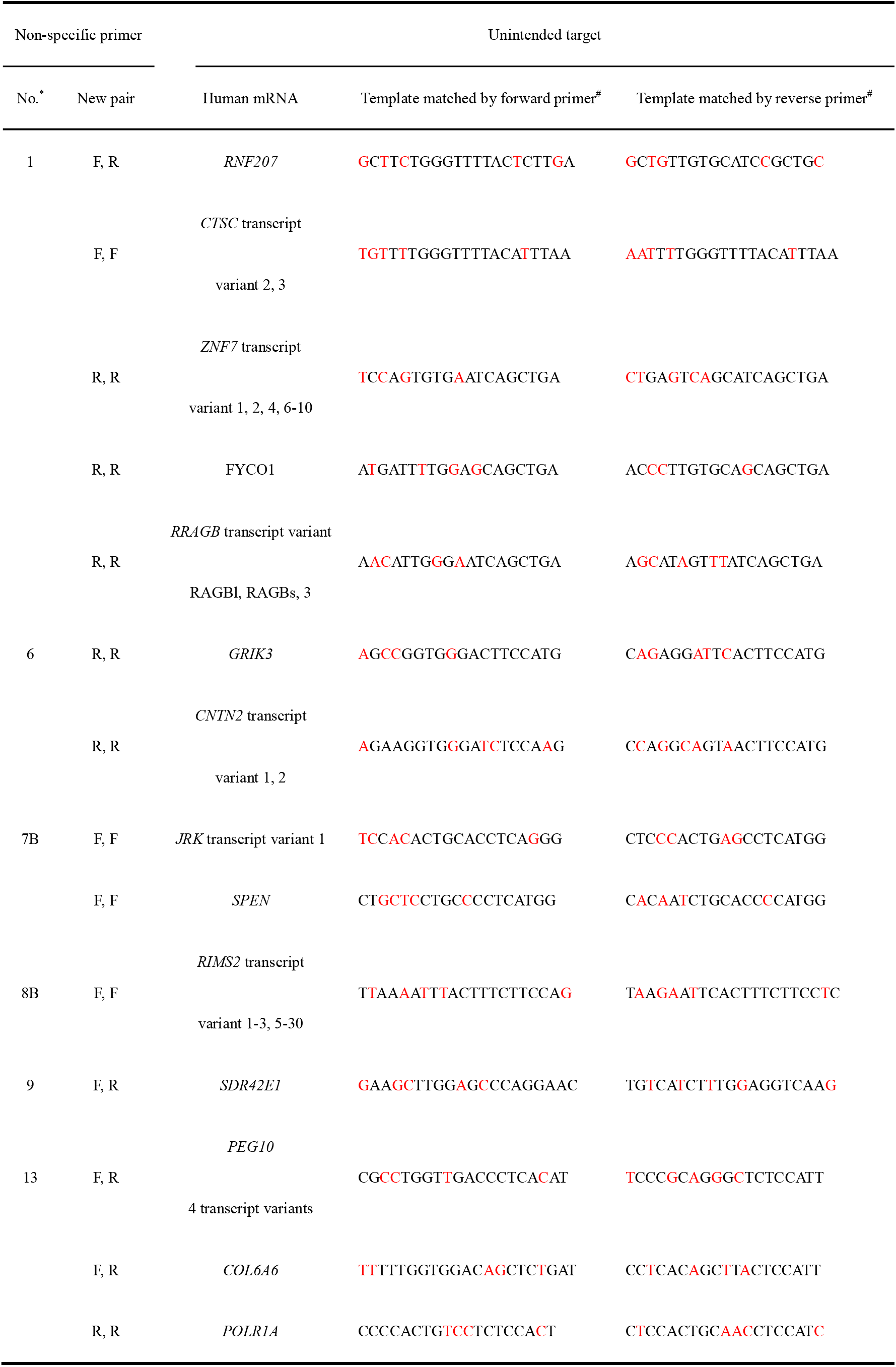
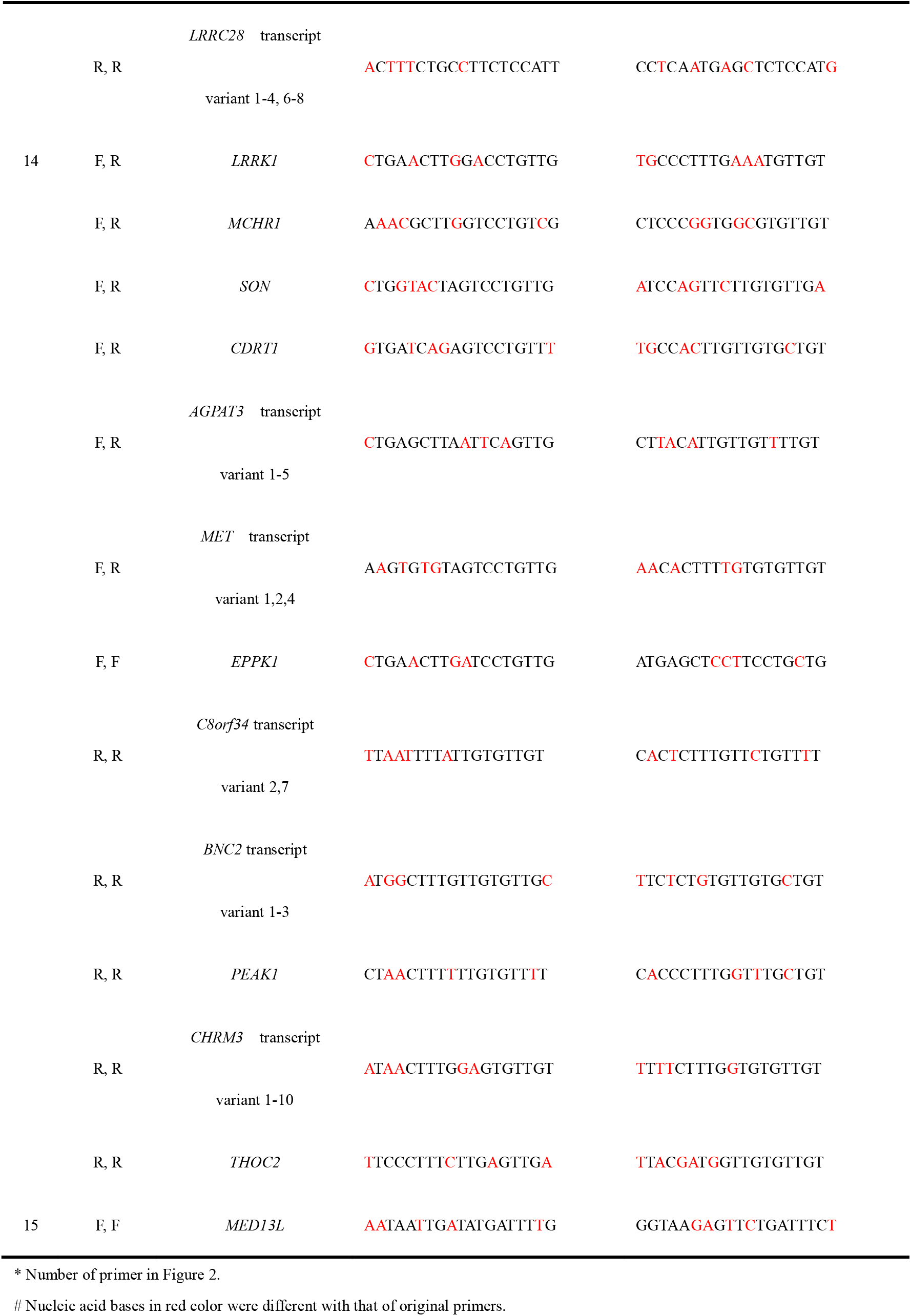
Non-specific primer and the unintended target.

Furthermore, all primers from NIID, MPH and IP and one primer from HKU in Table S1 had non-specific problems. Primer 6, 7B, 8B, 9, 13, 14 and 15 matched 3, 2, 29, 1, 13, 30 and 1 human mRNAs or their transcript variants, respectively. Although there were different clinical respiratory symptoms between SARS-CoV-2 and SARS virus, both of them belonged to beta coronavirus. Nevertheless, it was a remarkable fact that there were no any SARS primer pairs matched human mRNAs.^21^ Otherwise, it was noted that no pathogen mRNAs were considered except for human host.

Expression of human mRNAs targeted by primer 1 was shown in Figure 3 and those of other primers were also listed in Table S2. The result indicted that Cathepsin C (*CTSC*) mRNA had a much higher abundance than other mRNAs in the respective tissues, but difference of *CTSC* transcripts between individuals was not evident in tonsil. Relative to house-keeping gene glyceraldehyde-3-phosphate dehydrogenase (*GAPDH*), protein-transcripts per million (pTPM) values of *CTSC* mRNA in normal tonsil, lung and small intestine were 7.2%, 50.8% and 4.9% of *GAPDH* mRNA, respectively. The pTPM values gave a quantification of the mRNA abundance, which could be compared between different genes and specimens. Except for *CTSC, SON, THOC2* and *AGPAT3* transcripts, all other transcripts shown in Table S3 had smaller average pTPM values. It was noted that the latter three transcripts were targeted by same primer. Relative to house-keeping gene *GAPDH*, the total pTPM values of *SON, THOC2* and *AGPAT3* transcripts in normal tonsil, lung and small intestine were 6.5%, 18.0% and 8.3% of *GAPDH* mRNA, respectively.

**Figure 3:**
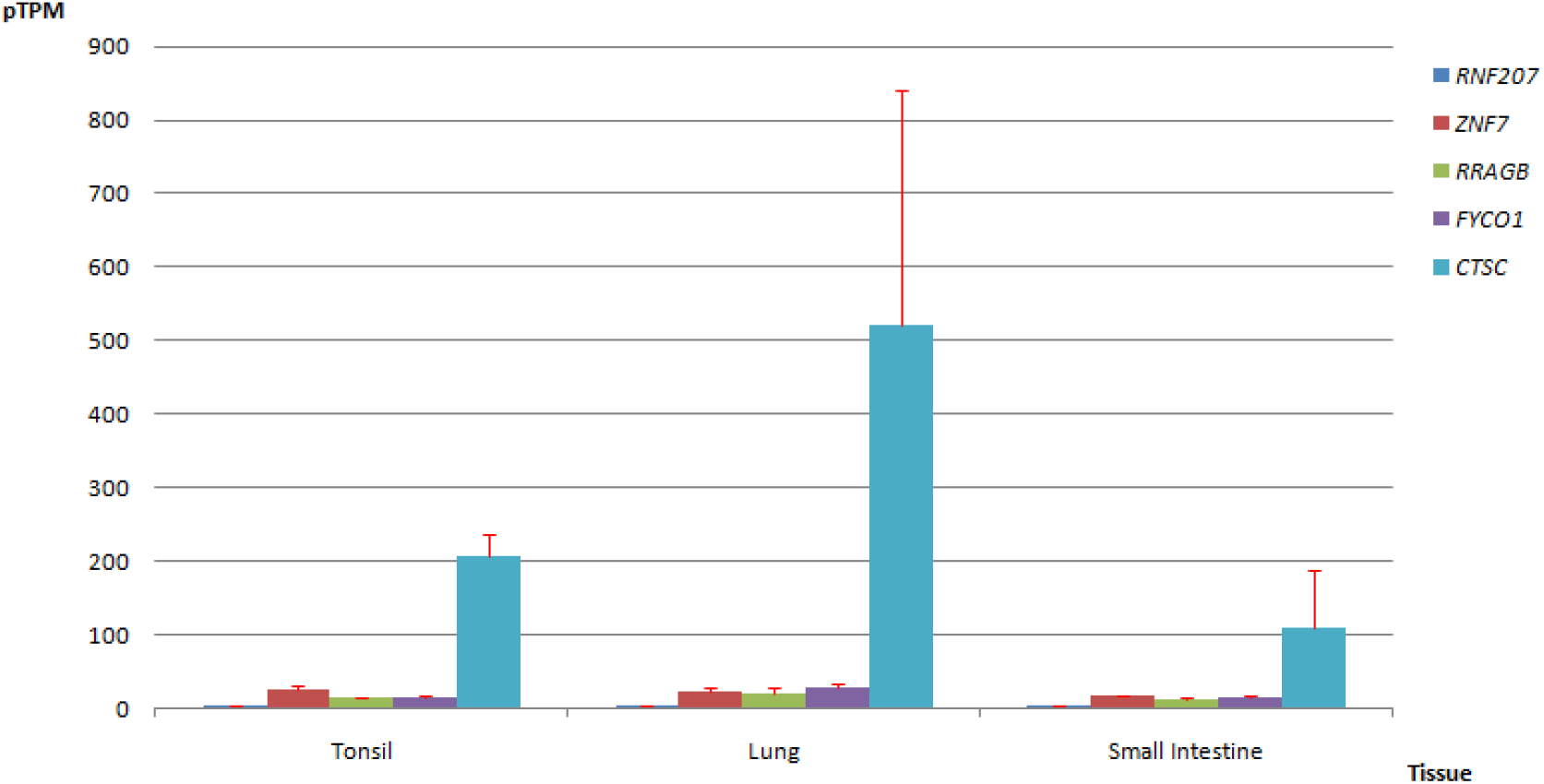
Expression of some target human mRNAs in different normal tissue.

## Discussion

Although many patients tested one or more negative before receiving positive results for SARS-CoV-2 virus in China, it was difficult to understand the extent to which this abnormal phenomenon occurs. However, the WHO reports on new laboratory-confirmed cases and new clinically diagnosed cases in China during the COVID-19 epidemic were helpful to deduce that extent. Rapid raise and drop of total new cases in Figure 1 indicated that the amazing number of clinically diagnosed cases was just an accumulation of those in Hubei province earlier than February 13, 2020. Successive revision in the diagnosis criterion for the novel coronavirus pneumonia in Hubei province had resulted in a sudden surge in the number of total new cases and the removal of clinically diagnosed cases in the province. ^16, 17^ There was no reports on whether the clinically diagnosed patients tested or not, but it was possible to infer general detection situation through evaluating the ability to detect COVID-19 infections. The ability to detect COVID-19 infections was steadily enhanced along with the outbreak of the epidemic and then exhibited a great accelerated growth. China National Biotec Group could produce novel coronavirus detection kits for 200,000 people each day on February 18, 2020.^18^ In the light of the linear fitting of laboratory-confirmed cases from January 22 to February 5, 2020 in the inset of Figure 1, the ability to detect COVID-19 infections for more than 14,400 persons besides laboratory-confirmed patients could be deduced from February 6 to February 13, 2020, and that basically met the requirements for 12,289 detections for clinically diagnosed patients on February 13, 2020. Hence it was inferred that a considerable number of clinically diagnosed patients could test and obtained negative results.

Moreover, the number of recovered cases in China moderately increased from 261 on February 5, 2020 to 3,622 on February 27, 2020, which was shown in Figure S1. It represented that cross-infection in hospital could be well controlled. Because there was no rapid increase of recovered cases after February 13, 2020, it could be concluded that most of more than 17,410 clinically diagnosed patients had finally diagnosed COVID-19 infections. The slowly decline of number of existing confirmed cases as shown in Figure S2 also supported the conclusion. Therefore, it was considered that false-negative results in RT-PCR detection of COVID-19 infections usually occurred among clinically diagnosed patients.

RT-PCR detection was applied to diagnose SARS virus in 2003, but little false-negative diagnosis was reported in more than 8000 confirmed cases. Furthermore, even if so many RT-PCR detections were repeated in laboratories and hospitals in China, general operation problems resulting in false-negative detections were not found. Consequently, the present study focused on the specificity of RT-PCR primers, that is, a pair of specific primers used to detect SARS-CoV-2 virus should only amplify the virus sequence, but not any unintended RNA, including those of human host and other pathogens. In contrast to pathogens, human host was usually omitted.

To eliminate the host interference in RT-PCR detection of COVID-19 infections, specific amplification of the intended target of SARS-Cov-2 sequence required that primers matched as little as possible to any human RNA transcript. Besides primer pairs matched *ZNF7, FYCO1* and *RRAGB* transcripts, it was noted that the primer pair with sixteen consecutive nucleotides at 3’ ends, which deeply matched those of *CTSC* transcripts except one nucleotide, could cause the serious interference during RT-PCR amplification. The technical guidance, including the primer pair 1 and 2, was employed to instruct centers for disease control at all levels in China.^6, 7, 9, 19, 20^ Although there was no interference with primer 2 for N gene, only both positive PT-PCR detection for COVID-19 infections with primer 1 and 2 were obtained in the same specimen could a positive result be confirmed in term of the clinical diagnostic criterion for nucleotide detection in China. Hence it was not difficult to draw a conclusion that non-specificity of the primer specifically designed for SARS-CoV-2 virus might be an important factor resulting in so many false-negative diagnoses for COVID-19 infections in China.

Moreover, there were seven other non-specific primer pairs. Although primer 7B and 8B were inner primers of nested RT-PCR, second round RT-PCR operation would be susceptible to contamination. Therefore, it was believed that false-negative diagnoses for COVID-19 infections might occur when using the above primers.

Besides the specificity of primers, the amount of the unintended target mRNA was another key factor in determining whether human host contamination could happen in RT-PCR detection. Method for sampling SARS-CoV-2 virus was similar to that for sampling SARS virus.^22^ RT-PCR detection specimens were collected usually from nasopharynx or oropharynx, sputum or fluid in trachea or alveoli and stool, which corresponded to tonsil, lung and intestine. Since viral particles, inflammatory cells and somatic cells mixed together in specimens and purification procedures could usually not separate viral RNA and host cellular during the RT-PCR detection, contamination from host RNA could not be avoided. The high abundance of *CTSC* transcripts in tonsil, lung and small intestine tissues and good match to primer 1 could lead to the contamination of clinical speciemens, which were used for RT-PCR detection of COVID-19 infections, and might trigger the false-negative diagnosis. Furthermore, the higher total abundance of *SON, THOC2* and *AGPAT3* transcripts in tonsil, lung and small intestine tissues and better matches to primer 14 could also result in the above problems. Although other transcripts had smaller pTPM values, expression of other transcripts might become very complex when inflammatory and oxidative stress occurred during viral infections.

According to the above mentioned analysis and data, there could be no doubt that non-specificity of RT-PCR primers obviously increased the risk of false-negative diagnoses for COVID-19 infections. Since there were so many false-negative diagnoses for COVID-19 infections in Hubei province at the early stage of epidemic and the reason was that some primers provided by institutes were nonspecific, Table 2 gave the suggestion on the RT-PCR primers in order to reduce false-negative results in RT-PCR detection for SARA-CoV-2 virus during COVID-19 pandemic.

**Table 2:**
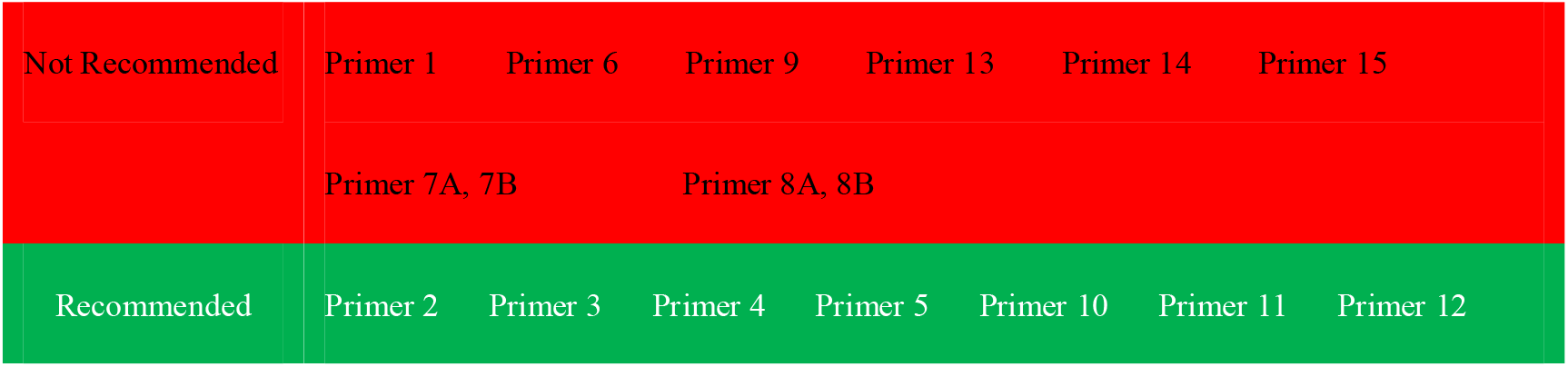
Suggestion on the RT-PCR primers for detection of SARA-CoV-2 virus.

## Data Availability

Numbers of new cases of COVID-19 infections were released by WHO. Seventeen specific primers for detection of Covid-19 were pbtained from WHO.All mRNA expressions in different human organs were obtained from Human Protein Atlas.

https://www.who.int/

https://www.proteinatlas.org/

## References

1. World Health Organization. WHO Director-General’s opening remarks at the media briefing on COVID-19-11 March 2020. Mar 11, 2020. https://www.who.int/dg/speeches/detail/who-director-general-s-opening-remarks-at-the-media-briefing-on-covid-19---11-march-2020 (accessed April 7, 2020).

2. Zhu N, Zhang D, Wang W, et al. A novel coronavirus from patients with pneumonia in China, 2019. N Engl J Med 2020; 382: 727–33.

3. World Health Organization. Coronavirus disease (COVID-2019) situation reports. April 15, 2020. https://www.who.int/docs/default-source/coronaviruse/situation-reports/20200404-sitrep-75-covid-19.pdf?sfvrsn=99251b2_4 (accessed April 7, 2020).

4. World Health Organization. Current list of WHO reference laboratories providing confirmatory testing for COVID-19. Mar 2, 2020.https://www.who.int/docs/default-source/coronaviruse/who-reference-laboratories-providing-confirmatory-testing-for-covid-19.pdf?sfvrsn=a03a01e6_2 (accessed April 7, 2020).

5. National Microbiology Data Center. Novel Coronavirus National Science and Technology Resource Service System. Jan 24, 2020. http://nmdc.cn/nCoV/en (accessed April 7, 2020).

6. Xie X, Zhong Z, Zhao W, Zheng C, Wang F, Liu J. Chest CT for typical 2019-nCoV pneumonia: relationship to negative RT-PCR testing. Radiology 2020; published online Feb 12. DOI: 10.1148/radiol.2020200343.

7. Zou L, Ruan F, Huang M, et al. SARS-CoV-2 viral load in upper respiratory specimens of infected patients. N Engl J Med 2020; published online Feb 19. DOI: 10.1056/NEJMc2001737.

8. Pan Y, Zhang D, Yang P, Poon LLM. Viral load of SARS-CoV-2 in clinical samples. Lancet Infect Dis 2020; published online Feb 24.DOI: 10.1016/S1473-3099(20)30113-4.

9. Lan L, Xu D, Ye, G, et al. Positive RT-PCR test results in patients recovered from COVID-19. JAMA 2020; published online Feb 27. DOI: 10.1001/jama.2020.2783.

10. Technical guidance for laboratory testing of 2019-nCoV infection (third edition). Biosafety and Health. published online Feb 5. DOI: 10.1016/j.bsheal.2020.02.001.

11. New coronavirus pneumonia prevention and control program (2nd edtion.) (in Chinese). Jan 22, 2020. http://www.nhc.gov.cn/jkj/s3577/202001/c67cfe29ecf1470e8c7fc47d3b751e88.shtml (accessed April 7, 2020).

12. Uhlen M, Oksvold P, Fagerberg L, et al. Towards a knowledge-based Human Protein Atlas. Nat Biotechnol 2010; 28(12):1248–50.

13. Ye J, Coulouris G, Zaretskaya I, Cutcutache I, Rozen S, Madden TL. Primer-BLAST: A tool to design target-specific primers for polymerase chain reaction. BMC Bioinformatics 2012; 13: 134.

14. Altschul SF, Gish W, Miller W, Myers EW, Lipman DJ. Basic local alignment search tool (BLAST). J Mol Biol 1990; 215(3): 403–410.

15. Shi H, Han X, Jiang N, et al. Radiological findings from 81 patients with COVID-19 pneumonia in Wuhan, China: a descriptive study. Lancet Infect Dis 2020; published online Feb 24. DOI: 10.1016/S1473-3099(20)30086-4.

16. New coronavirus pneumonia prevention and control program (5th edtion.) (in Chinese). Feb 21, 2020. http://www.nhc.gov.cn/jkj/s3577/202002/a5d6f7b8c48c451c87dba14889b30147.shtml (accessed April 7, 2020).

17. New coronavirus pneumonia prevention and control program (6th edtion.) (in Chinese). Mar 7, 2020. http://www.nhc.gov.cn/jkj/s3577/202003/4856d5b0458141fa9f376853224d41d7.shtml (accessed April 7, 2020).

18. The State Council of the People’s Republic of China. China’s central SOEs mobilize to fight against coronavirus. Feb 19, 2020. http://english.www.gov.cn/statecouncil/ministries/202002/19/content_WS5e4c9d14c6d0595e03c210a0. html (accessed April 7, 2020).

19. Wang D, Hu B, Hu C, et al. Clinical characteristics of 138 hospitalized patients with 2019 novel coronavirus-infected pneumonia in Wuhan, China. JAMA 2020; published online Feb 7. DOI: 10.1001/jama.2020.1585.

20. Guan W, Ni Z, Hu Y, et al. Clinical characteristics of coronavirus disease 2019 in China. N Engl J Med 2020; published online Feb 28. DOI: 10.1056/NEJMoa2002032.

21. World Health Organization. PCR primers for SARS developed by WHO network laboratories. April 17, 2003. https://www.who.int/csr/sars/primers/en/ (accessed April 7, 2020).

22. World Health Organization. Sampling for Severe Acute Respiratory Syndrome (SARS) diagnostic tests. April 29, 2003. https://www.who.int/csr/sars/sampling/en/ (accessed April 7, 2020).

